# Is the end near? When the different countries will surmount COVID-19 pandemic: new approach applying physical, mathematical and game theory models

**DOI:** 10.1101/2020.12.01.20242099

**Authors:** J. G. García de Alcañíz, V. López-Rodas, E. Costas

**Affiliations:** Genetics, Faculty of Veterinary Medicine, Complutense University, 28040 Madrid, Spain

**Keywords:** COVID-19 duration, Delta-*t* argument, Cooperate strategy, Defect Strategy, central limit theorem, binomial probability

## Abstract

In the year 2020 COVID-19 pandemic was a global issue that changed mankinds lifestyle. Since then, when we will control the pandemic and recover our normal life has become the paramount question to be answered, and it needs to be solved. One problem is that there are wealthy countries, with very good health care systems and scientific resources while others barely dedicate 100 US $ per citizen per year, rich countries could cooperate at different levels with poorer ones. In such a diverse context classic epidemiology models, excellent for predicting short term evolution of the pandemic at a local level are not as suitable for long term predictions at a global scale specially if the data they use are of questionable accuracy. Alternatively, big data and AI approaches have been tried. There is an option that can be more effective. Physics applies predictive models about the duration of an event based on analysing the dynamics of the time evolution of the event itself. These models can be used alongside with probabilistic and game theory models that consider different degrees of cooperation. By means of the physics Delta-*t* argument and a game theory model (cooperate versus defector) we calculate when different countries may control COVID-19 pandemic. In a non-cooperate model, those countries with more resources and best manage the pandemic will have it under control between May and September 2021, whereas those with no resources will suffer the pandemic until at least October 2023. On the other hand, a strong cooperative model will allow that the majority could control the COVID-19 pandemic between October 2021 and November 2022.

## Introduction

Last days of 2019 saw the first diagnosed cases of a novel coronavirus in Wuhan city, China^1^. Soon it jumped from a local outbreak to a world scale problem, causing the pandemic that has brought the world to its knees. Many countries have failed in their attempt to control the spread of SARS-CoV-2 through preventive measures (social distancing, increased hygiene, use of masks, etc.) and immense use of resources and expense. To date, the Covid-19 pandemic has produced more than 62.662.181 infected and over 1.460.223 deaths. The pandemic is also taking its toll from the world’s economy, according to the International Monetary Fund, the economic activity is likely to remain subdued until health risks abate^2^.

Nonetheless, since the outbreak, our comprehension of the pandemic and the virus has been slowly growing (i.e., scientific evidence of aerosol transmission, importance of the use of face masks, better medical procedures, increased efficiency in early detection and follow up programs, etc.) allowing us a better response to this global crisis.

Mankind’s greatest hope rests on being able to develop an effective vaccine that can be made available, as soon as possible, to thousands of millions of people. According to Krammer, it is likely that vaccination will require more than one dose, this means that at least 16 billion doses should be needed to meet the global demand^3^. Worldwide, the scientific community, pharmaceutical companies and governments are engaged in a common quest for a vaccine, using huge amounts of public and private resources in many countries on an unprecedented scale.

Under normal circumstances, fifteen years to complete a vaccine development is not uncommon^3^. Only once a vaccine was developed in a record time of four years. Measles virus was isolated 1963 and a vaccine from that strain (Jeryl Lynn strain) was approved for its use in 1967^4,5^

The graveness of this Covid-19 pandemic has led to an unknown situation in history. The world is in a race to obtain a vaccine, cutting down usual completion times is paramount. With the use of vaccination programs, as Gregg, A. in 1949 pointed out, the incidence of many diseases has been moved from the area of chance to the area of choice^6^. Pharmaceutical companies have made great progress and are ready to start with the vaccination programs COVID-19 pandemic may now be entering the area of choice, although still in the area of chance some light may be seen ahead.

The big question -yet to be answered- is when vaccines, anti-SARS-CoV-2 drugs, epidemiology strategies and increased herd immunity will put an end to the pandemic.

Since there are natural wildlife reservoirs^7–9^ and in many people it produces an asymptomatic infection^10–13^, many virologists think that SARS-CoV-2 is here to stay. However, it is very different to have a virus that is not under control and unleashes a global pandemic with millions of infected and dead, to one that can just be a seasonal occurrence and is not a mayor threat.

Here we propose a nouvelle mathematical approach that may help answer afore mentioned big question. It may give some clues as when the different countries will definitively control SARS-CoV-2.

Classic predictions about the pandemic, based on epidemiological models, are unreliable due to the lack of knowledge, Will vaccination programs work? Will natural or vaccine-induced immunity last? Will SARS-CoV-2 mutate to avoid vaccination efforts? These are just a few of the numerous unanswered questions, as there has not been a similar situation in our history. There are many concerns about what the future may bring.

In these circumstances, we consider that some elegant mathematical approaches, based on basic science, physics and probability principles could shed light over the future that lays ahead, when countries will start winning the battle against the virus.

Specifically, we consider that a Copernican approach, the Gott’s Delta-*t* argument is of great help under the current set of events. It proves especially useful when predicting complex phenomena with a great deal of uncertainty, as the moment when different countries will control SARS-CoV-2 is. Delta-*t* argument has been used before to predict Human race’s longevity or when the Berlin wall would fall^14^. This delta-*t* argument has already been used to predict the Covid-19 pandemic duration and the number of infected and dead by SARS-CoV-2^15^.

With Delta-*t* argument we can know the time interval in which COVID-19 pandemic will be under control. It is clear that not all countries will reach that moment at the same time, some will do it sooner and for others it may take longer. The time interval obtained using delta-*t* argument has a lower limit, corresponding to those countries that will control the pandemic earlier, and an upper limit, that will correspond to those last countries controlling COVID-19 pandemic. Countries within the lower limit will have applied better epidemiological strategies (social distancing, wide use of face masks, massive detection tests, trackers, …), will have better health care systems, better and sufficient drugs, enough vaccine doses, effective massive vaccination programs, etc. Countries that need more time to control the COVID-19 pandemic will rely more on herd immunity.

But there are other mathematical procedures that allow an estimate of how the time distribution of the different countries controlling COVID-19 pandemic will be.

There are two procedures that permit it:

One of these procedures assumes that all countries behave as totally independent entities. There is no cooperation between them, and if one achieves the goal of controlling SARS-CoV-2 it will not affect the speed at which others will defeat the virus.

The other model has the opposite view, a country controlling COVID-19 pandemic increases the likelihood of another doing the same. In case of the COVID-19 pandemic we believe the world works closer to this second model. Even though one particular country develops a vaccine, it will not be only used in its nationals, it will be distributed to many other countries as well, shortening the time period needed to have COVID-19 pandemic under control worldwide.

Using both predictive models can be interesting in the future. A comparison of the expected results with these theoretical models and the observed results when the COVID-19 pandemic is under control can be used to evaluate the efficiency of international cooperation.

## Material and Methods

### Theoretical background

#### Delta-targument

Estimating when countries will control the virus using epidemiological arguments is difficult, but by using the Delta-*t* argument we can predict the most likely time interval when this can happen.

A very detailed explanation of the application of the Delta-*t* argument to calculate the most probable duration of the Covid-19 pandemic is detailed in García de Alcañiz et al (2020)^15^. The problem of the application of the Delta-*t* argument to estimate when countries will curb the pandemic with a given probability is similar, in short:

Considering our position as not privileged, Copernican principle^14,16^ we can predict the availability in time of countries containing the disease. It is clear that any observable event can only be measured between its commencement time (*t*_*begin*_) and its final time (*t*_*end*_), if we are in a non-privilege moment nor in space or time, we can assume that our current time (*t*_*now*_) will be randomly placed across the duration of the event. For so, ratio *r*=(*t*_*now*_−*t*_*begin*_)/(*t*_*end*_−*t*_*now*_)has to be a random number between 0 and 01. In this way, we can calculate the probability of any future event, in our study countries controlling the pandemic.

**Figure 1.**
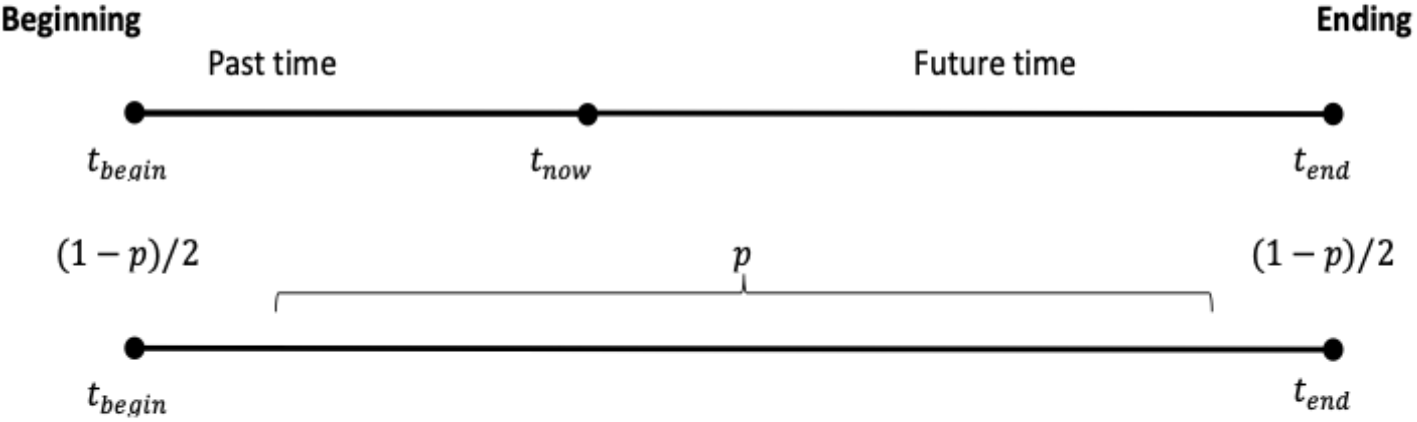
time distribution of a non-privileged event

Considering afore mentioned premisses, the different countries controlling the COVID-19 pandemic and its related probability can be calculated using equation 1.

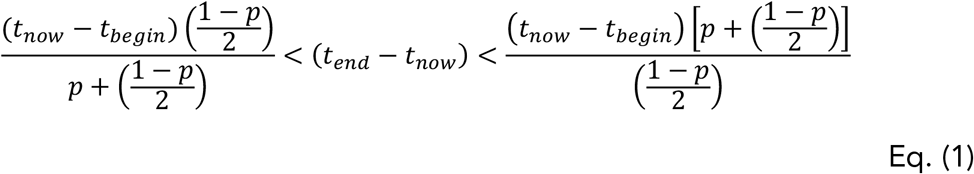

Countries that sooner suppress COVID-19 pandemic will do it in a time period closer to that estimated by the first term of the Eq. (1) inequality. Whereas those countries needing more time to do it will be closer to the estimated time given by the third term of the Eq. (1) inequality.

#### COVID-19 pandemic lifetime in the different countries

With the Delta-*t* argument we can estimate the time interval in which the COViD-19 pandemic will be under control.

It is of great interest to estimate how the time distribution of dates when countries control COVID-19 pandemic will be. In our opinion it is very different to have most countries grouped at an early stage or have them at the end of the time period estimated by the Delta-*t* argument.

This can be also predicted using probabilistic arguments and game theory. Two theoretical opposite models can occur:

- <u>Model 1.- Defect Strategy</u> (pandemic duration if countries do not cooperate).

The premise is that countries behave independently, without cooperation at all. In this scenario, what happens to one country does not affect the events on any other country. In our case, moment at which different countries control COVID-19 pandemic. With this setting we can estimate the time distribution of these periods using the central limit theorem and the cumulative distribution function.

The central limit theorem establishes that, when taking sufficiently large (usually n > 30) random independent samples under common conditions, the sum of many random variables (such as: the moment when countries will control the pandemic) will have an approximately normal distribution (reviewed in Zabell 1995, Jorgensen 1997, Fisher 2010, Montgomery & Douglas 2014)^17–20^. Consequently, the temporal distribution of the frequencies of the dates at which countries will control SARS-CoV-2 will follow a normal probability distribution in the time interval within the limits obtained using Delta-*t* argument -i.e. lower limit be the earliest countries controlling the COVID-19 pandemic and upper limit the latest countries doing so- (to wit a classic statistical problem demonstrated decades ago, Yule & Kendall 1950, Fisher 1954, Cochran & Cox 1957, Sokal & Rohlf 1959)^21–24^. Using the cumulative distribution function in this special case of a standard normal distribution Φ(*z*), we can estimate the probability of any interval as:

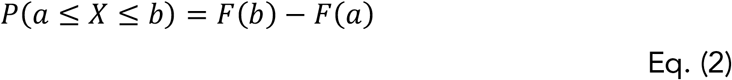

where *P*(*a*≤*X*≤*b*)is the probability of controlling the pandemic in the interval (*ab*). The different countries will end with COVID-19 pandemic following temporal a distribution characterized by the central limit theorem. The probability of controlling the pandemic before a certain date can be estimated using the cumulative distribution function:

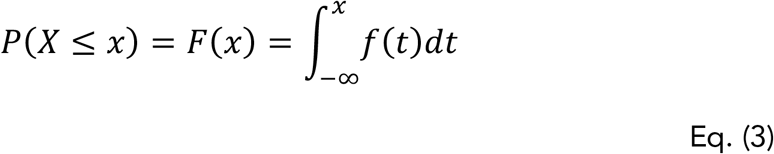

The number of countries that will have COVID-19 under control before a certain date is obtained by multiplying the total number of countries by their probability.

- <u>Model 2. Cooperate strategy</u> (Pandemic duration if countries cooperate).

In their fight against SARS-CoV-2 countries do not work independently. On the contrary, if one country controls COVID-19 pandemic favours other countries controlling the disease as well because they share vaccines, drugs and successful epidemiological strategies. This would put COVID-19 pandemic to an end sooner than expected than if every country worked independently.

If a cooperative strategy model is taken to its maximum theoretical limit of cooperation, all countries would end COVID-19 exactly at the same time. The moment in time when this could happen would be close to the earliest possible moment predicted by Delta-*t* argument.

However, reality will be different because some countries will better manage strategies against SARS-CoV-2 than others, for instance by having more resources and logistics to implement massive vaccination programs.

To make this cooperate strategy model adjust better to reality we will classify selected countries into four categories:

One group of countries, capable of developing their own drugs and vaccines and to implement successful control strategies without needing aid from other countries. A second group of countries, that cannot develop vaccines or drugs against SARS-CoV-2 on time but have enough resources to buy and use them efficiently. A third group of countries that rely on international cooperation to obtain drugs or vaccines but has the necessary infrastructure to reasonably use them. Finally, a group of countries with no logistics to even use efficiently international aid.

Countries with less health care resources and poorer logistics will need more time to control COVID-19 despite the international aid.

With this model of cooperate strategy we can estimate the time distribution of dates at which the different countries will be free of COVID-19, using a binomial probability distribution^25,26^ as follows:

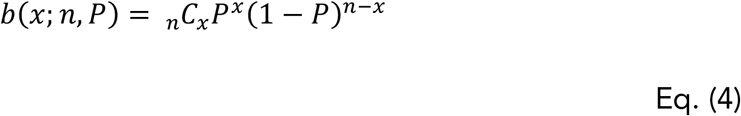

Where: *b* =binomial probability; *x* =total number of “successes” (pass or fail, heads or tails etc.); *P* =probability of a success on an individual trial; *n* =number of trials

### Practical Background

It would not be unreasonable to think that SARS-CoV-2 will stay among human populations for decades since it can infect asymptomatically to many people and has natural reservoirs in animals.

However, sometime in the future COVID-19 will no longer be a pandemic. We assume in our model that one country is free from SARS-CoV-2 when there are isolated outbreaks as a rare event incapable to increase its frequency in the population (i.e., rates under 1 per million people)

For our study we selected the same countries selected by García de Alcañiz et al (2020) in his paper^15^ plus all those countries from the World Health Organisation’s web page that showed more than a thousand cases throughout the pandemic, this brings a total figure of 172 countries.

In Model 2 (cooperate strategy), we consider that those countries that allocate less than 100 US dollars per person for health care will not have the logistics needed to make the most of the international collaboration. Its considered that 41,7% of world’s countries are in this situation.

## Results and discussion

Usually, when approaching a new problem, we rely on tools or methodologies that have been tested and proven to be useful. This has been the case with COVID-19 pandemic, scientists and epidemiologists around the globe have been using traditional epidemiology tools but at a local scale(i.e. SIR, SIRD models^27–29^, Gompertz’s equation^30–35^, etc.). However, skepticism arises when using these tools with COVID-19 pandemic at a global scale because data used to feed the models are also unreliable. Number of infected and dead, R values, infectivity rates, mortality rates, etc., create a lot of uncertainty due to its great variability among countries (different strategies to control the pandemic, different methodologies to process data, reliability of official data, etc.). All these make results from traditional epidemiology models not as useful to analyse COVID-19 pandemic at a global scale as it may seem at first, these factors hamper the relevance and reliability of these traditional models^15^.

Other science disciplines, to analyse extremely complex phenomena and produce useful results, have come to use other mathematical approaches not common to epidemiology. Some of these elegant mathematical approaches, based on basic science, physics and probability principles, are the Copernican principle and the Delta-*t* argument, Lindy’s Law, the Doomsday principle-Carter’s catastrophe, all of which allow predicting complex phenomena characterized by their great uncertainty, as the Covid-19 pandemic is^15^. Regardless of the use of these reductionist physic approaches in a series of seminal works (i.e. Thomson, 1917; Schrödinger, 1944; Morowitz, 1970; Lima de Faria, 1988; Margulis & Sagan, 1995)^36–40^ that enabled spectacular advances in biology and medicine, especially in molecular aspects, these physic-mathematical approaches are unfortunately scarcely used in epidemiology.

In the present work, we use physic-mathematical models to estimate how long the COVID-19 pandemic will last in the different countries.

In our model we use boundary conditions that allow us to estimate the earliest possible date at which countries can control the COVID-19 pandemic. We also take on two opposite strategies that the different countries can follow: One would be what we called the defect strategy, by which every country tackles the pandemic alone. The second possibility is what we named as cooperate strategy, where all countries work cooperatively.

### Delta-*t* argument

Delta-*t* argument to calculate the duration of the COVID-19 pandemic outcome is a time interval between to ends:

At one end will be the earliest possible date at which the COVID-19 pandemic could be over. This will only happen in an ideal country that followed all successful epidemiology strategies (widely use of face masks, social distancing, confinement policies, early detection with massive PCRs, maximum contacts tracking, etc.), with the best health care system, excellent health care professionals, enough and effective anti-SARS-CoV-2 drugs, enough and effective vaccines and with the logistics to vaccinate quickly and massively all the population.

In the other end of the time interval is the latest possible date to get over COVID-19 pandemic. That end logically corresponds to those countries with no efficient management of the pandemic, no health care system or a very poor one, with not enough drugs or vaccines and not even the logistics to successfully administer them. In this extreme setting COVID-19 will be self-controlled when herd immunity is reached, or the number of less harmful SARS-CoV-2 strains dominate in the population.

In order to calculate this interval, we need to establish two boundary conditions for the Delta-*t* argument. The accuracy of the prediction will hinge on these boundary conditions.

One boundary condition is the (*t*_now_ –*t*_begin_) parameter, we consider this to be of twelve months. Although possible that SARS-CoV-2 was present in human populations at an earlier date, it was around November 2019 when the first cases started to be detected in Wuhan city in China^1^.

The other one is the *p* value. Defining *p* value, type 1 and 2 errors in which we can fall into have to be considered. If we give a high *p* value the time interval of the prediction increases a lot, up to the point of being useless. For *p* =01the time interval would go from the present moment to the infinite future. In line to those who use Delta-*t* argument we will define a *p* value of *p* =0, 5^14,16,41–44^.

Delta-*t* argument predicts that some countries with better resources and strategies to manage COVID-19 pandemic will commence to be out from April 2021 onwards. On the other hand, countries with worst management and poorer resources will not succeed until the end of 2023.

- <u>Model 1.- Defect strategy</u> (COVID-19 pandemic duration if countries do not cooperate)

However, Delta-*t* argument says nothing about how many countries will free themselves from COVID-19 at a certain date, but this estimate is of great interest. It is quite different that a majority of countries are free from COVID-19 by summer 2021 or by the end of 2023.

Combining Delta-*t* argument with central limit theorem we can have an estimate of the number of countries that will be free from COVID-19 at a certain date (Table 1).

**Table 1.** COVID-19 pandemic duration if countries follow a defect strategy. First row represents time intervals; second one represents the probability of ending the pandemic in that time interval; third row is the number of countries that will be free of the pandemic in that time interval.

| **Time interval** | Until Apr. 30<sup>th</sup>, 2021 | From May 1<sup>st</sup> to Sep. 30<sup>th</sup>, 2021 | From Oct. 1<sup>st</sup>, 2021 to Feb. 28<sup>th</sup>, 2022 | From Mar. 1<sup>st</sup>, to Jul. 31<sup>st</sup>, 2022 | From Aug. 1<sup>st</sup>, to Dec. 31<sup>st</sup>, 2022 | From Jan. 1<sup>st</sup>, to May 31<sup>st</sup>, 2023 | From Jun. 1<sup>st</sup>, to Oct. 31<sup>st</sup>, 2023 | After Nov. 1<sup>st</sup>, 2023 |
| --- | --- | --- | --- | --- | --- | --- | --- | --- |
| **Probability** | 0.0014 | 0.0213 | 0.1360 | 0.3413 | 0.3413 | 0.1360 | 0.0213 | 0.00014 |
| **Nº. of countries** | 0 | 4 | 23 | 59 | 59 | 23 | 4 | 0 |

Those countries that better manage the pandemic will start to be out of it by the summer of 2021. The majority will be out between the winter and autumn of 2022. Some will suffer the COVID-19 pandemic until the autumn or winter 2023.

Nevertheless, it is important to remember that the assumption of this model is that all countries behave as independent entities, without cooperation. One country getting over COVID-19 pandemic does not affect the moment others will follow.

However, if countries cooperate results would be different.

- <u>Model 2.- Cooperate strategy</u> (pandemic duration if countries cooperate)

According to Delta-*t* argument, from April 2021 some countries that better manage the COVID-19 pandemic with better resources, enough vaccines and drugs against SARS-CoV-2 may be overcoming the pandemic.

In a cooperative model, when one country is over the pandemic it will increase the likelihood of other countries following.

We combine the outcome of Delta-*t* argument with a cooperate strategy model with four different country types (i.-countries capable of developing vaccines and with effective control strategies; ii.-countries that do not develop vaccines but have enough resources to buy them and means to use them efficiently; iii.-countries that cannot buy vaccines but do have infrastructure to efficiently apply any international aid they may get; iv.-countries without logistics to efficiently use international aid), and considering boundary conditions that assure that the model produces the best possible result (i.e. minimum time needed for the different country types to control COVID-19 pandemic).

Predictions obtained with this model are summarised in Table 2.

**Table 2.** COVID-19 pandemic duration if countries follow cooperate strategy. First row represents time interval. Second row shows the probability of pandemic ending at each time interval. Third row shows the number of countries overcoming the pandemic on each time interval.

| **Time interval** | From May 1<sup>st</sup>, 2021 until Sep. 30<sup>th</sup>, 2021 | From Oct. 1<sup>st</sup>, 2021 to Apr. 30<sup>th</sup>, 2022 | From May 1<sup>st</sup>, 2022 to Oct. 31<sup>st</sup>, 2022 | After Nov. 1<sup>st</sup>, 2022 |
| --- | --- | --- | --- | --- |
| **Probability** | 0.1971 | 0.4248 | 0.3051 | 0.0073 |
| **No. of countries** | 34 | 73 | 52 | 1 |

Countries that better manage COVID-19 pandemic and with enough resources will start to be out from May 2021 and will have the pandemic under control by September 2021. An important group of countries will end with COVID-19 between the autumn of 2021 and the spring of 2022. Countries more dependent on international aid will not be capable of controlling the pandemic until the autumn of 2022. It may happen that, last one out of COVID-19 pandemic will be much longer than that date.

It is necessary to highlight that this model shows the best possible theoretical scenario, reproducing the minimum time needed for the different countries to go back to normality.

Comparison of both models -defect and cooperate strategy-show the significance of international cooperation. A strong cooperative model would have COVID-19 pandemic reasonably under control by November 2022 allowing to return to normal life worldwide. Without that cooperation another year will be needed It is clear that COVID-19 pandemic is not only a medical or scientific problem, it has economic, politic and social implications that make predictions very unstable and in continuous change.

## Data Availability

All data is of public access and was retrieved from oficial web pages

